# Non-Invasive markers of inflammation and protein loss augment diagnosis of celiac disease

**DOI:** 10.1101/2023.05.24.23290489

**Authors:** Kimberly A Sutton, Mai He, Changqing Ma, Ta-Chiang Liu, William A. Faubion, Julie Hoffman, Laura Linneman, Cynthia Rodriguez, Lori R Holtz

## Abstract

**Background:** Circulating tissue transglutaminase IgA (TTG IgA) concentrations are sensitive and specific indicators of celiac disease, but discrepancies between serologic and histologic findings still occur. We hypothesized that fecal markers of inflammation and protein loss would be greater in patients with untreated celiac disease than in healthy controls. Our study aims to evaluate multiple fecal and plasma markers in celiac disease and correlate these findings with serologic and histologic findings as non-invasive means of evaluating disease activity.

**Methods:** Participants with positive celiac serologies and controls with negative celiac serologies were enrolled at the time of upper endoscopy. Blood, stool and duodenal biopsies were collected. Concentrations of fecal lipocalin-2, calprotectin and alpha-1-antitrypsin and plasma lipcalin-2 were determined. Biopsies underwent modified Marsh scoring. Significance was tested between cases and controls, modified Marsh score and TTG IgA concentration.

**Results:** Lipocalin-2 was significantly elevated in the stool (*p*=0.007) but not the plasma of participants with positive celiac serologies compared to controls. There was no significant difference in fecal calprotectin or alpha-1 antitrypsin between participants with positive celiac serologies and controls. Fecal alpha-1 antitrypsin >100mg/dL was specific, but not sensitive for biopsy proven celiac disease.

**Conclusions:** Lipocalin-2 is elevated in the stool but not the plasma of patients with celiac disease suggesting a role in the local inflammatory response. Calprotectin was not a useful marker in the diagnosis of celiac disease and did not correlate with degree of histologic changes on biopsy. While random fecal alpha-1 antitrypsin was not significantly elevated in cases compared to controls, an elevation of greater than 100mg/dL was 90% specific for biopsy proven celiac disease.

## Introduction

Celiac disease is an immune mediated enteropathy triggered by gluten in the diet and mediated by antigen presenting cells, intraepithelial lymphocytes (IEL) and T cells (1). The standard screening test for celiac disease is circulating tissue transglutaminase IgA (TTG IgA). TTG IgA is highly sensitive and specific (2), but mismatch between serological and histological findings still exist, resulting in diagnostic uncertainty. Additionally, while TTG IgA is used for monitoring response and adherence to gluten free diet, it is not a reliable predictor of mucosal healing (3). These limitations of TTG IgA could be minimized with additional biomarkers.

Fecal markers can assess inflammation of the gut. Fecal calprotectin is a neutrophil marker of inflammation commonly used in the screening of inflammatory bowel disease (IBD) and monitoring disease activity (4). The value of fecal calprotectin in diagnosis of celiac disease has been variably established (5-7).

Fecal neutrophil gelatinase-associated lipocalin (lipocalin-2) is an acute phase reactant expressed by several cell types including neutrophils and enterocytes (8). Fecal lipocalin-2 concentration is elevated in patients with active disease and decreases with mucosal healing in patients with IBD (9, 10). Fecal lipocalin-2 has comparable sensitivity and specificity to fecal calprotectin in its ability to predict clinical and endoscopic disease activity in IBD (11). Additionally, the expression of *LCN2*, the gene that encodes lipocalin-2, is increased in the duodenum of patients with environmental enteric dysfunction, another small bowel enteropathy with histopathologic similarities to celiac disease (12, 13). A prior study found no elevation of plasma lipocalin-2 in patients with celiac disease (14). While immunohistochemistry for lipocalin-2 shows staining in small bowel biopsies of children with celiac disease and not in healthy controls, a difference was not found on quantification (13). We are not aware of any data regarding fecal lipocalin-2 in celiac disease.

Fecal alpha-1 antitrypsin (A1AT) concentration, which indicates protein leakage from the circulation into the gut, is increased in people with celiac disease, and correlates with disease activity (15). Protein loss in celiac disease occurs due to structural change in the tight junctions leading to disruption of the function of the epithelial barrier (16).

We hypothesized that markers of inflammation and protein loss would be elevated in patients with newly diagnosed, treatment-naive celiac disease compared to healthy controls. Our study aims to evaluate multiple markers in celiac disease and correlate with serologic and histologic findings as a non-invasive means of evaluating disease activity.

## Methods

### Participants

This study was approved by the Washington University in St. Louis Human Research Protection Office. Informed consent was obtained from the parent(s), and assent from children 12 years old or older. Cases were defined as children ages 0-17.9 years with abnormal celiac serologies (TTG IgA, TTG IgG, deamidated gliadin IgA, deamidated gliadin IgG, or endomysial ab), on a gluten containing diet who were referred for upper endoscopy. Patients with a history of biopsy proven celiac disease were excluded. Controls were defined as children ages 0-17.9 years with normal celiac serologies. Patients with a history of known celiac disease or other inflammatory disorders of the gut were excluded.

### Celiac Serologies

Serologies were often obtained before referral to GI clinic and performed by different clinical laboratories with varying reference ranges based on the assay used. TTG IgA results were normalized by relating to the upper limit of normal of each results’ respective reference range.

### Sample Collection

Stool samples were collected at home by participants using a stool collection kit provided by the research team. Samples were transported on ice and stored at -80° C. Plasma samples were collected on the day of the upper endoscopy, transported on ice, and stored at -80° C.

### Biopsies

Biopsies were obtained from the duodenal bulb (2 pieces) and the second portion of the duodenum (4 pieces), per our center’s protocol (17). Two pathologists blinded to the participants serology results reviewed each biopsy and assigned a modified Marsh score (18). If scores were concordant, this was considered the final score. If the scores diverged by one step (e.g. 3a and 3b), then the higher score was used. If the scores diverged by more than one step, a third blinded pathologist assigned a modified Marsh score. If two of the three scores agreed, that score was used. If all three scores were different, the middle score was used.

### Lipocalin-2, calprotectin and alpha-1 antitrypsin concentrations

Plasma and stool lipocalin-2 concentrations were determined using a commercially available kit to perform a sandwich ELISA for human lipocalin-2 (Human Lipocalin-2/NGAL R&D Systems, Minneapolis, MN, USA). Plasma samples were diluted 1:1000 using the reagent diluent buffer provided in the kit. Fecal samples were diluted 1:10,000 by serial dilution. The first dilution of 1:50 used fecal extraction buffer (0.1 M Tris. HCL (8.0), 0.15 M NaCl, 1 M urea, 10mM CaCl2, 0.1 M citric acid monohydrate, 5g/l BSA and 0.25 mM Thimerosal). Subsequent dilutions used the reagent diluent buffer provided in the kit.

Fecal calprotectin concentrations were determined using a commercially available kit to perform a sandwich ELISA for human calprotectin (Human Calprotectin HycultBiotech, Wayne, PA, USA). Samples were diluted 1:5000 by serial dilution.

Fecal A1AT concentrations were determined using a commercially available kit to perform a sandwich ELISA for fecal alpha-1 antitrypsin (Alpha-1 Antitrypsin Immunochrom, Heppenheim, Germany). Samples were diluted 1:12,500 by serial dilution.

For each assay, a standard curve was done on every plate and samples were run in duplicate. Coefficient of variation was less than 15% for each duplicate.

### Statistics

Significance was tested between two groups using Mann-Whitney test and Fisher’s exact test for continuous and categorical variables, respectively. Linear regression was used for continuous variables. Kruskal-Wallis test with post-hoc testing was used to determine significance across groups. Receiver operating characteristic (ROC) analysis was used to determine sensitivity and specificity. All p-values were two sided, and α values <0.05 were considered statistically significant. Statistical analyses were performed using Prism 9.3.1 (GraphPad).

## Results

### Demographics

In total, 73 cases and 18 controls were included in this study. The median age was 7 years old for cases and 13 years old for controls with 70% girls and 30% boys (table 1). Conditions with increased risk of celiac disease were represented in our cases, but not our controls. Specifically, among the cases there were 5 patients with trisomy 21, 9 patients with type one diabetes mellitus, 1 patient with hypothyroidism and 1 patient with Turner syndrome.

**Table 1.** Demographics

|  | Cases (n=73) | Controls (n= 18) | p |
| --- | --- | --- | --- |
| Girls (%) | 51 (70) | 13 (72) | ns |
| Median age years (IQR) | 7 (5-11) | 13 (10-16) | <.0001 |
| Race Caucasian (%) | 72 (99) | 18 (100) | ns |
| Ethnicity Hispanic (%) | 1 (1) | 0 | ns |
| Trisomy 21 (%) | 5 (7) | 0 | ns |
| Type 1 DM (%) | 9 (12) | 0 | ns |
| Hypothyroidism (%) | 1 (1) | 0 | ns |
| Turner Syndrome (%) | 1 (1) | 0 | ns |

### Histopathology

Of the controls, one patient had *Helicobacter pylori* infection in gastric biopsy and one had eosinophilic esophagitis. No other diagnoses were made based on histopathology in the control participants.

The modified Marsh score agreed within 1 level for 62 participants. A third pathologist was used for 11 scores to resolve score differences. Among the cases, 20 were Marsh 0, 1 was Marsh 1, and 52 were ≥ Marsh 3a.

### Serology

Five cases were referred for endoscopy for elevation of other celiac serologies with normal TTG IgA (Table 2). Of those 5, 1 case had IgA deficiency. The degree of TTG IgA elevation did not correlate significantly with modified Marsh score among cases who were referred for endoscopy for elevated TTG IgA. Among cases with elevated TTG, there was significant difference in degree of fold change of TTG IgA to upper limit elevation when comparing Marsh groups 0-1 with Marsh 3a-c combined *p*=0.009).

**Table 2.** Cases Serology Results

|  | Positive TTG IgA (n = 68) |  | Negative TTG IgA (n = 5) |  |
| --- | --- | --- | --- | --- |
|  | Negative | Positive | Negative | Positive |
| TTG IgG | 3 | 6 | 0 | 4 |
| Deamidated Gliadin IgA | 3 | 6 | 1 | 0 |
| Deamidated Gliadin IgG | 0 | 4 | 0 | 1 |
| Endomysial IgA | 3 | 21 | 1 | 0 |

### Plasma and Stool Markers

We measured the plasma concentration of lipocalin-2 and concentrations of lipocalin-2, calprotectin and A1AT in stools (Figure 1). Fecal lipocalin-2 was significantly higher in stool from cases than controls (p = 0.0074) (Figure 1B). We found that concentrations of fecal lipocalin-2 differed by modified Marsh score (p = 0.0367) (Figure 2B). We combined Marsh 3a-c to test if fecal lipocalin-2 could detect a difference between those with biopsy evidence of celiac disease and controls, and again found significantly higher concentrations of fecal lipocalin-2 in participants with celiac disease compared to controls (*p* = 0.0095) (Supplementary figure 1A). Among those with elevated serology, we compared those with Marsh 3a-c to Marsh 0 and found no significant difference in fecal lipocalin-2 concentrations (Supplementary figure 1B). Plasma lipocalin-2 concentration was comparable between cases and controls (Figures 1A) and could not distinguish degree of histopathology based on modified Marsh score (Figure 2A).

**Figure 1.**
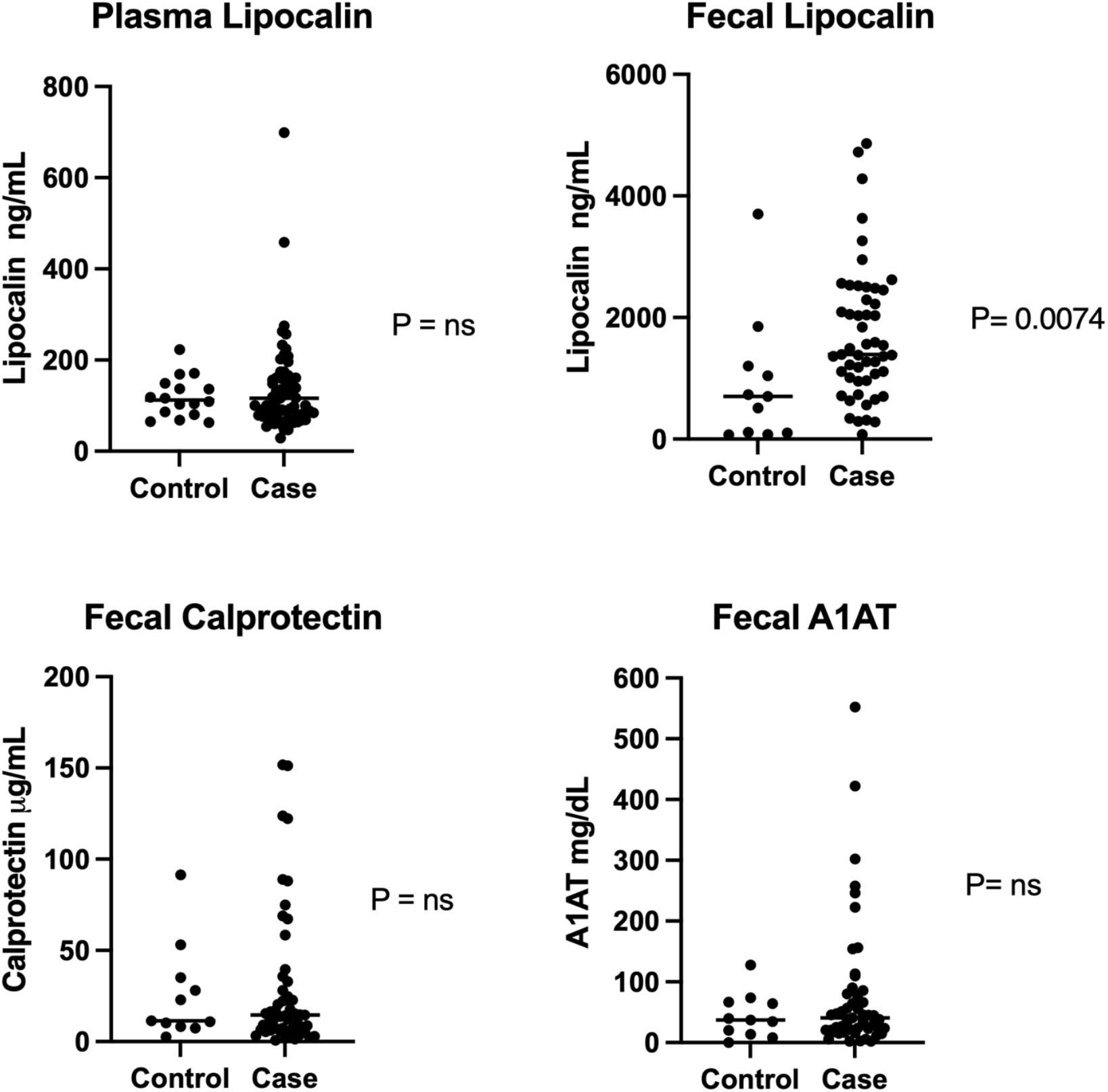
Comparison of fecal and plasma markers in cases vs. controls. P values from Mann-Whitney test. (A) Plasma lipocalin-2 results. (B) Fecal lipocalin-2 results. (C) Fecal Calprotectin results. (D) Fecal A1AT results. A1AT: Alpha-1 Antitrypsin. Horizontal lines represent medians.

**Figure 2.**
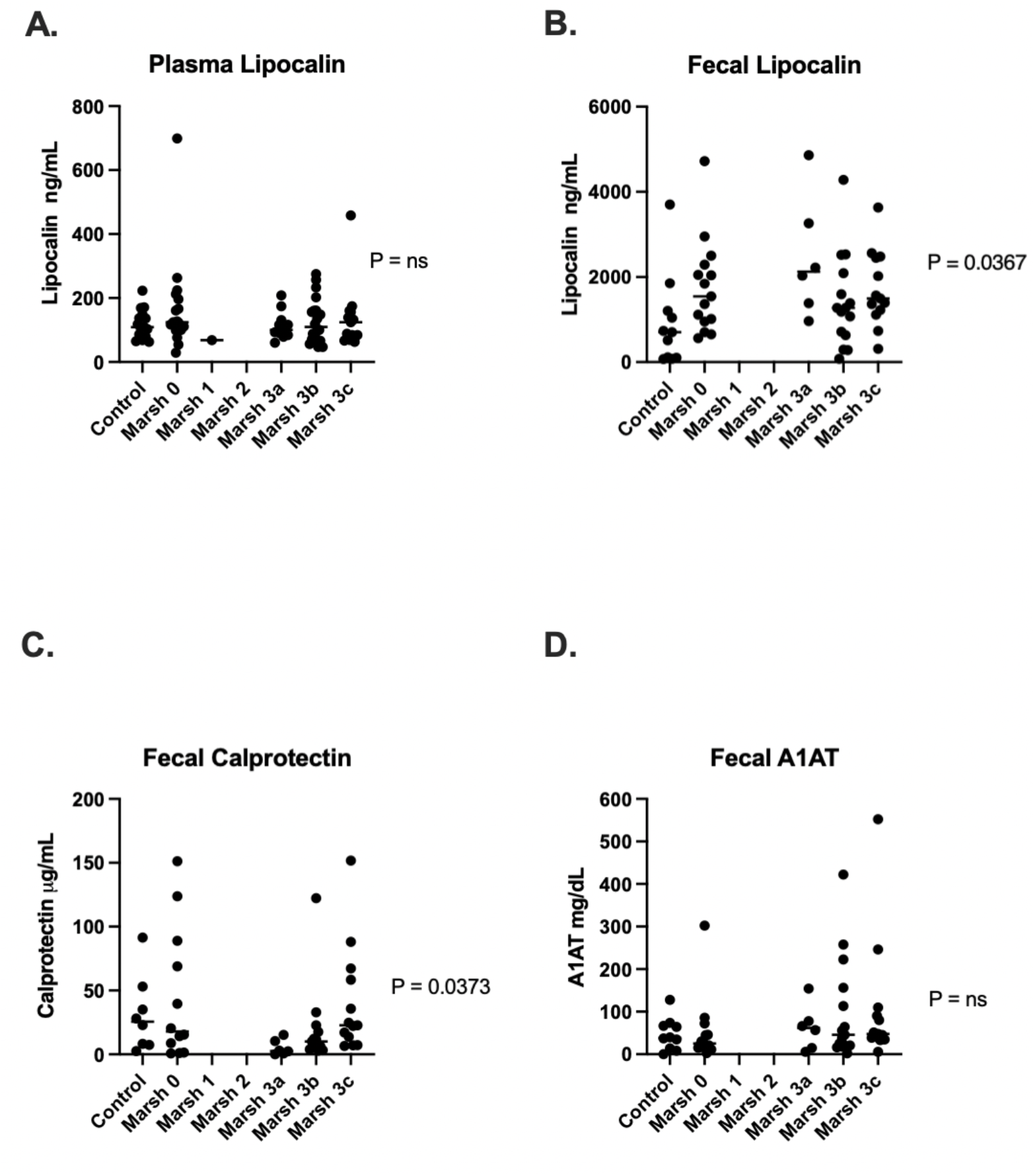
Comparison of fecal and plasma markers vs modified Marsh score. P values from Kruskal-Wallis test. (A) Plasma lipocalin results. (B) Fecal lipocalin-2 results. (C) Fecal calprotectin results. (D) Fecal A1AT results. A1AT: alpha-1 antitrypsin. Horizontal lines represent medians.

Fecal calprotectin concentrations did not significantly differ in stools between cases and controls (Figure 1C), but showed significant variation when stratified by the degree of duodenal mucosal damage (*p* =0.0373) (Figure 2C). However, on post-hoc analysis of the groups, only the medians of the Marsh 3a and Marsh 3c groups were significantly different (*p* = 0.0373), and there was no significant difference between any of the Marsh 3 groups and either Marsh 0 or controls. Combining Marsh 3a-c to test if fecal calprotectin could detect a difference between those with biopsy evidence of celiac disease and controls, also showed no difference (Supplementary Figure 2).

Fecal A1AT concentrations did not differ between cases and controls (Figure 1D) or by degree of histopathology (Figure 2D). However, we noted that a fecal A1AT concentration greater than 100 mg/dL which is the threshold for clinically significant protein losing enteropathy, had a specificity of 91% for distinguishing cases versus controls, but only a sensitivity of 20%. The specificity of a fecal alpha-1 antitrypsin of >100 mg/dL for biopsy confirmed disease (defined as a modified Marsh score ≥3a or greater) was 90% compared to controls and 93% compared to cases with elevated serologies but normal biopsies (Marsh 0) with sensitivity of 26% for each.

To determine if fecal markers or plasma lipocalin-2 differed with the degree of circulating TTG IgA concentrations, we examined cases with elevated TTG IGA and found no significant association for any of the markers with the degree of TTG IGA elevation (Figure 3).

**Figure 3.**
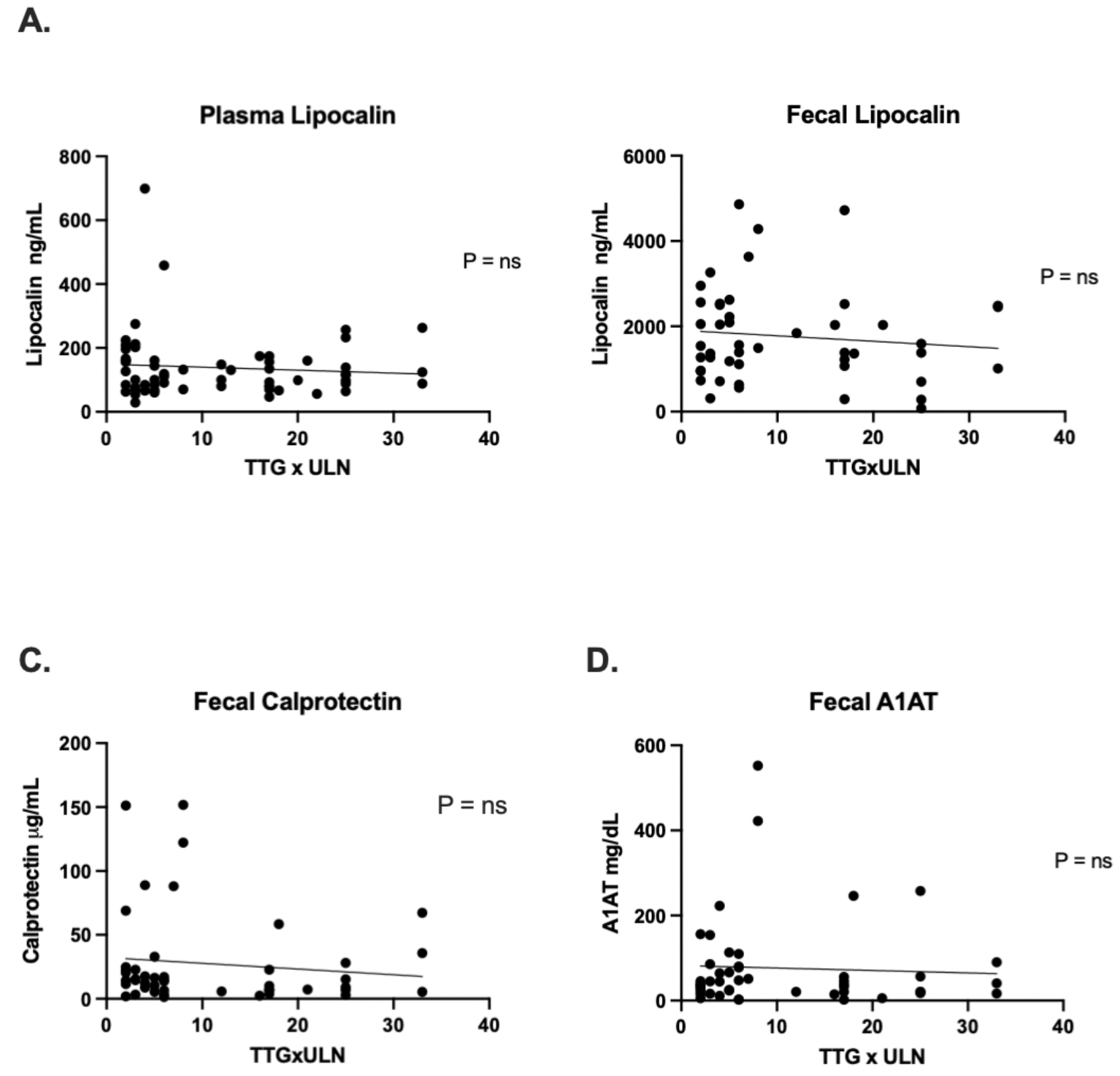
Comparison of fecal and plasma markers vs normalized TTG. P values from Linear Regression. (A) Plasma lipocalin-2 results. (B) Fecal lipocalin-2 results. (C) Fecal calprotectin results. (D) Fecal A1AT results. A1AT: Alpha-1 Antitrypsin. ULN: upper limit of normal

## Discussion

The lack of plasma elevation of lipocalin-2 agrees with the findings of Janas et. al (14). The difference in the stool and plasma findings suggests a difference in the local intestinal inflammatory response or epithelial turnover compared with the systemic inflammatory response. As lipocalin-2 is produced by both immune cells as well as the enterocytes themselves, either could be the source of the lipocalin-2 found in the stool.

Fecal lipocalin-2 differentiated children with biopsy confirmed celiac disease as defined by a modified Marsh score of 3a or greater from controls. However, it was not able to differentiate cases with biopsy confirmed celiac from cases who were seropositive but had non-confirmatory biopsies of Marsh 0. This suggests that the Marsh 0 cases may have some degree of intestinal inflammation which could represent early celiac disease or potentially have another inflammatory process leading to non-specific elevation of celiac serologies.

No difference in fecal calprotectin was found between cases and controls, further supporting recent European Society for Pediatric Gastroenterology, Hepatology and Nutrition (ESPGHAN) consensus that calprotectin is not a useful marker in celiac disease diagnosis based on mixed results of prior studies (19). We did find a significant difference between the medians of the Marsh 3a and 3c groups but without a difference compared to either the controls or the Marsh 0 group this is unlikely to have clinical relevance.

We did not find a significant elevation of fecal A1AT in cases compared with controls. Previous studies have found a significant increase in A1AT clearance in patients with active celiac disease (15). The lack of difference in our cohort may be due to the use of spot testing versus clearance. While random stool testing is easier for patients in the clinical setting, it does not account for circulating concentrations of A1AT and variation within stools samples which decreases the sensitivity of the test compared with 24 hour fecal A1AT clearance tests (20).

While the median A1AT of the cases was not significantly elevated compared to controls, there was a trend towards higher A1AT concentrations in the cases (Figure 1D) which we investigated further with ROC analysis. The clinical laboratory cutoff for a protein losing enteropathy is 100mg/dL. This cutoff had good specificity in our cohort for biopsy confirmed celiac disease, but poor sensitivity. We have found that a normal random A1AT does not rule out celiac disease, but a level elevated in the range of clinically significant protein losing enteropathy could distinguish patients with positive serologies when comparing with controls. Further, a random fecal A1AT >100mg/dL was predictive of participants that had biopsy confirmed disease compared with participants that had positive serologies but non-confirmatory biopsies.

A potential limitation of this study may be the selection of controls who underwent upper endoscopy for gastrointestinal symptoms, though no duodenal inflammation was found on their biopsies. Another potential limitation is the difference in the median ages between the case and controls groups. While the cases represent the age range of new onset celiac disease patients in our center, the controls skewed older which likely represents the typical age of functional abdominal pain presentation as those are the patients most likely to undergo upper endoscopy without known gastrointestinal inflammation and negative celiac serologies. While the comorbidities of trisomy 21, type one diabetes, hypothyroidism, and Turner syndrome were not represented in the control group, this is representative of the increased risk of celiac disease in patients with these disorders.

In conclusion, we have found elevation of lipocalin-2 in stool, but not plasma, to be a marker of in patients with elevated celiac serologies. Calprotectin was not useful in diagnosing celiac disease. Clinically significant elevations of A1AT in the stool is specific but not sensitive for biopsy confirmed celiac disease when compared with controls. Further studies are needed to investigate the change in fecal lipocalin-2 concentrations in response to gluten free diet as well as compared with mucosal healing after treatment. Our data endorse studying further fecal biomarkers to confirm or refute presence of celiac disease in ambiguous diagnostic situations.

## Data Availability

All data produced in the present study are available upon reasonable request to the authors

---

**Supplemental Figure 1.**
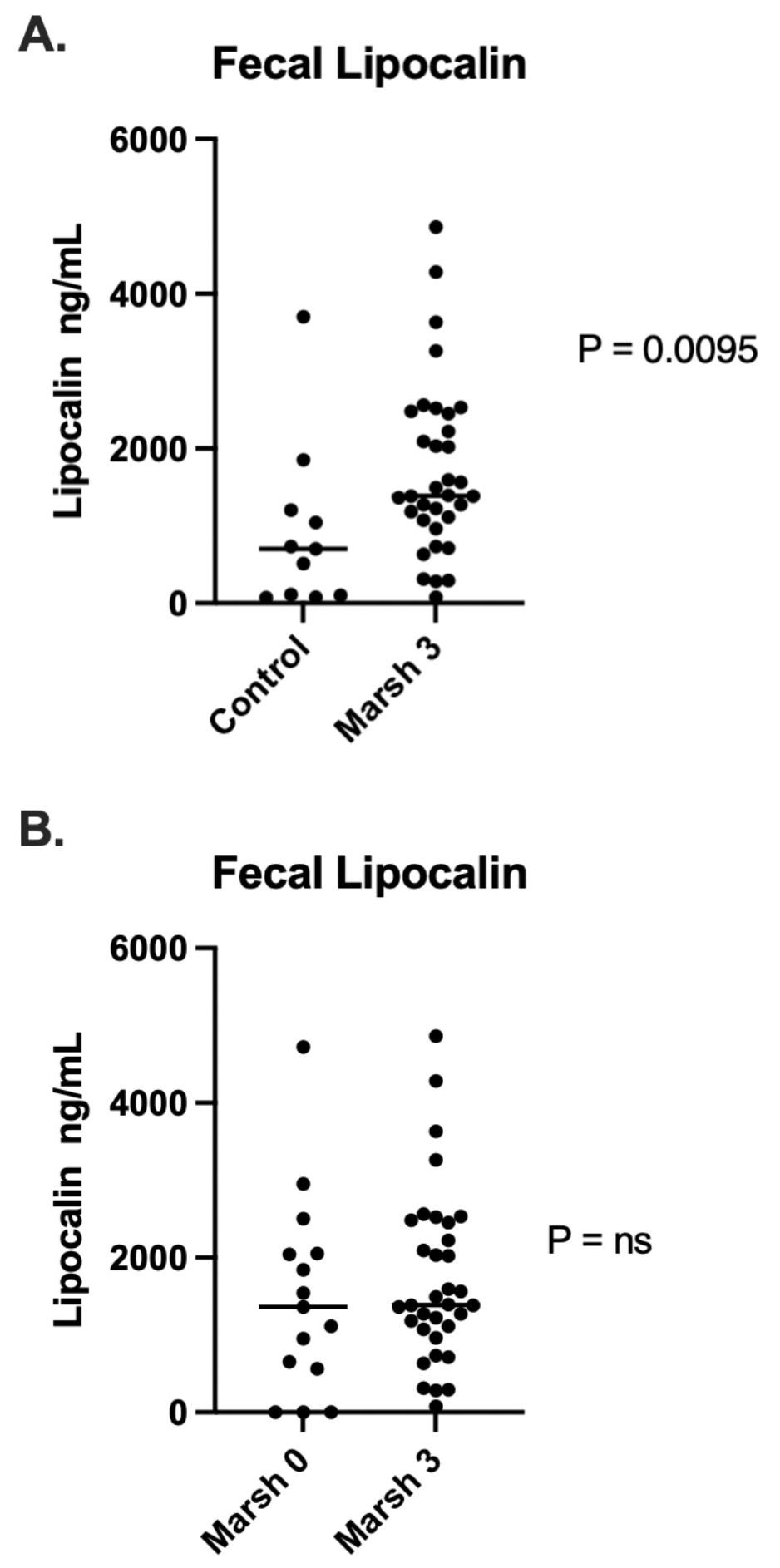
Comparison of fecal lipocalin-2 in biopsy confirmed vs biopsy negative patients and controls. P values from Mann-Whitney. (A) Fecal lipocalin-2 Marsh 3 compared to controls. (B) Fecal lipocalin-2 Marsh 3 compared to Marsh 0. Horizontal lines represent medians.

**Supplemental Figure 2.**
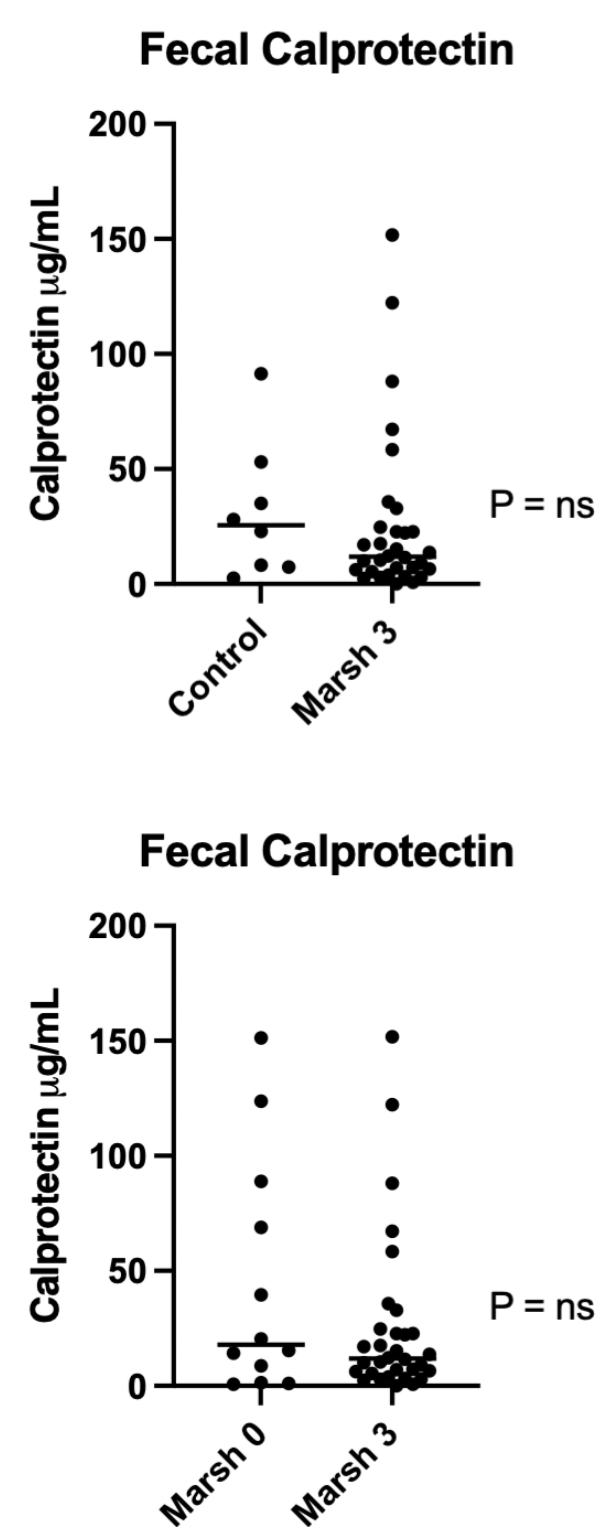
Comparison of Fecal Calprotectin in biopsy confirmed vs biopsy negative patients and controls. P values from Mann-Whitney. (A) Fecal Calprotectin Marsh 3 compared to controls. (B) Fecal Calprotectin Marsh 3 compared to Marsh 0. Horizontal lines represent medians.

